# The impact of patient body mass index on surgeon posture during simulated laparoscopy

**DOI:** 10.1101/2020.11.24.20237123

**Authors:** Ryan Sers, Steph Forrester, Massimiliano Zecca, Stephen Ward, Esther Moss

## Abstract

Laparoscopy is a cornerstone of modern surgical care. Despite clear advantages for the patients, it has been associated with inducing upper body musculoskeletal disorders amongst surgeons due to the propensity of non-neutral postures. Furthermore, there is a perception that patients with obesity exacerbate these factors. Therefore, novice, intermediate and expert surgeon upper body posture was objectively quantified using inertial measurement units and the LUBA ergonomic framework was used to assess the subsequent postural data during laparoscopic training on patient models that simulated BMI’s of 20, 30, 40 and 50 kg/m^2^. In all experience groups, the posture of the upper body significantly worsened during simulated surgery on the BMI 50 kg/m^2^ model as compared to on the baseline BMI model of 20 kg/m^2^. These findings suggest that performing laparoscopic surgery on patients with severe obesity increases the prevalence of non-neutral upper body posture and may further increase the risk of musculoskeletal disorders in surgeons.

## Introduction

Over the past thirty years Laparoscopic Surgery (LS) has revolutionised patient care and has quickly become the default interventional procedure within a myriad of surgical specialties [1–4]. The transition from open surgery has been supported by shorter recovery periods, less postoperative pain and lower risk of operative complications for the patient [5,6].

Despite the clear advantages of LS for the patient, the shift away from open surgery seems to have had a negative impact on surgeon health; specifically, an increased prevalence of musculoskeletal disorders (MSDs) [1,7]. Laparoscopic surgery, as compared to open surgery, has been associated with a significantly greater risk of MSDs in the neck, thorax and shoulders [8], with MSD complaints reported in 88% of 244 surgeons [9]. The most likely cause of increased rates of MSDs is the increase in non-neutral postures adopted by surgeons during LS [10–12].

The emergence and impact of patients with obesity (BMI ≥ 30 kg/m^2^) is an issue that has been linked with further deterioration of surgeon posture during LS [13]. Obesity classification is subdivided into three categories: Class 1, 30 kg/m^2^ < BMI ≤ 35 kg/m^2^, Class 2, 35 kg/m^2^ < BMI ≤ 40 kg/m^2^ and Class 3, BMI > 40 kg/m^2^ often termed as ‘severe’[14]. Obesity incidence in the UK has doubled in the past two decades with 29% of the adult population having obesity and 4% severely obesity [15]. A similar trend is observed worldwide [16].

Laparoscopic surgery is recognised as the optimal interventional technique for many obesity related pathologies. While literature suggests that Class 1 - 2 obesity has a minimal impact on surgeon posture during LS [13,17], there is mounting evidence that indicates patients with Class 3 obesity have a sub-optimal impact on surgeon posture, promoting non-neutral postures in the problem areas previously raised [9,18–20]. Moreover, an impending shortage of experienced surgeons would mean greater patient workloads per surgeon and less experienced surgeons within the workforce, which has the potential to further increase the prevalence of MSDs in surgeons [21]. The reliance on LS, rising obesity prevalence and an impending shortage of experienced surgeons prompts the query of what impact does patient body mass index (BMI) have on developing laparoscopists. Advanced trainees and non-expert surgeons are likely to be exposed to patients with high BMIs earlier and more frequently in their careers, therefore, understanding how obesity interacts with surgical experience is paramount. The primary aim of this study was to quantify the impact of rising patient BMI on surgeon posture, with a secondary aim of identifying how patient BMI interacts with surgical experience in affecting surgeon posture. The authors hypothesized that increased patient BMI would degrade surgeon posture and that this degradation would be greatest for less experienced surgeons.

## Materials and Methods

### Participants and Ethics

This study was conducted at the Sports Technology Institute at Loughborough University from November 2018 to June 2019. Ethical clearance for this study was attained from the Loughborough University Ethics Approvals Sub-Committee and all participants provided voluntary informed consent before testing commenced. Study participants included four university medical students familiar with simulated training environments but with no live LS experience (referred to as ‘novices’), three senior gynaecological trainees with 4 - 6 years’ LS experience (referred to as ‘intermediates’ with > 100 laparoscopic procedures a year) and three senior gynaecological surgeons with 10+ years of LS experience (referred to as ‘experts’ with > 100 laparoscopic procedures a year).

### Instrumentation and Equipment

The Perception Neuron V2 inertial motion capture system (NOITOM Ltd, China) was setup in the 18-neuron mode as validated in [22,23], and previously used within [24,25]. Only the upper body portion of the motion capture system was used within this study and consisted of 11 ‘physical’ (Table 1) and 4 ‘virtual’ neurons. Each ‘physical’ neuron consisted of an inertial measurement unit (IMU) with the following specifications: 3-axis accelerometer (± 16g), 3-axis gyroscope (± 2000 dps), 3-axis magnetometer, dimensions of 12.5 mm x 13.1 mm x 4.3 mm in size and a mass of 1.2 grams. Each ‘virtual neuron’ provided orientation data calculated by the systems proprietary algorithm giving data at the neck and approximately the T3, T8 and L1 vertebrae (Figure 1). All physical IMUs were connected in series to a wireless hub with dimensions of 59 mm x 41 mm x 23 mm. Motion data for all neurons was transferred via TCP/IP (120 Hz) to an external PC and imported into MATLAB 2019b (MATLAB, MathWorks, Natick, MA, USA) for analysis.

**Table 1.** Physical IMU positions [32].

| Physical sensor/neuron number<br>(Phys within Figure 1 and Table 2) | Anatomical position |
| --- | --- |
| 1 | Head |
| 2 | Upper thorax (C7) |
| 3 & 4 | Acromion (L & R) |
| 5 & 6 | Centre of humerus (L & R) |
| 7 & 8 | Proximal end of ulna (L & R) |
| 9 & 10 | Dorsal side of hand (L & R) |
| 11 | Lower thorax |

**Figure 1.**
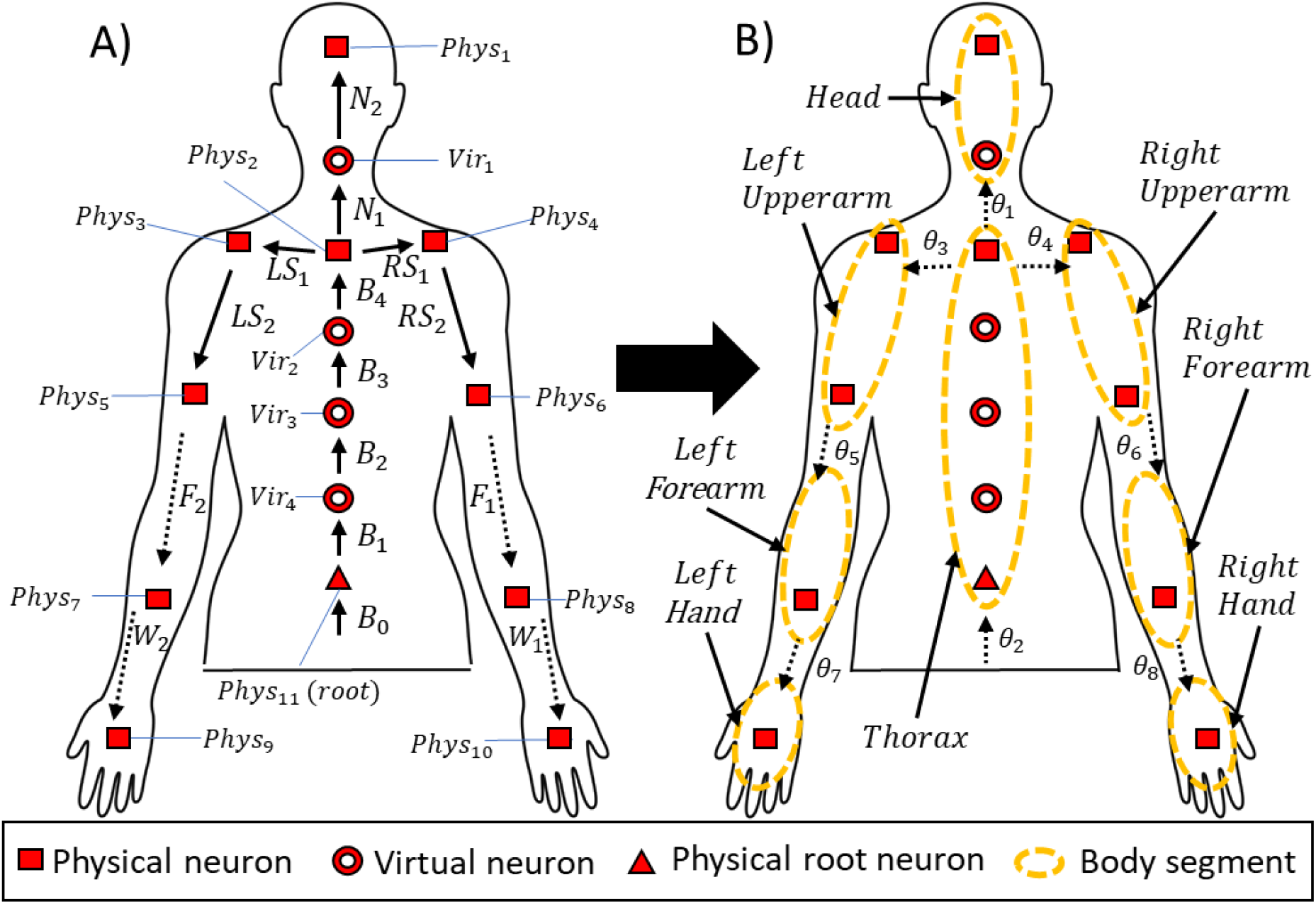
(A) Representation of the kinematic chains for the Perception Neuron model and (B) the adapted model illustrates the joint/segment angles used in the study. In (A) *B*_0−4_ denote thorax quaternion 4D vectors and *N*_1−2_, *LS*_1−2_, *RS*_1−2_, *F*_1−2_, *W*_1−2_ denote neck, shoulder, elbow and wrist quaternion vectors retrospectively. In (B) the dotted circles illustrate the new rigid body segments where the quaternion outputs from the enclosed neurons in each circle were combined through quaternion multiplication following an identical process defined in [23]. This method ensures that a single quaternion output represents the orientation of each body segment θ_1−8_ when defined at the joint which conjoins the child and parent segments (Table 2).

Participant segment lengths were obtained from images captured by an Xbox One Kinect 2 (Xbox, Microsoft, Redmond, WA, USA) and quantified using bespoke software written in MATLAB. The suit was then calibrated following the developers recommended process [26]. In addition, three static calibrations were taken with the participant in an I pose were taken; the participant stood with their hands parallel to their sides to facilitate a neutral posture. This acted as the baseline reference for all measurements.

**Table 2.**
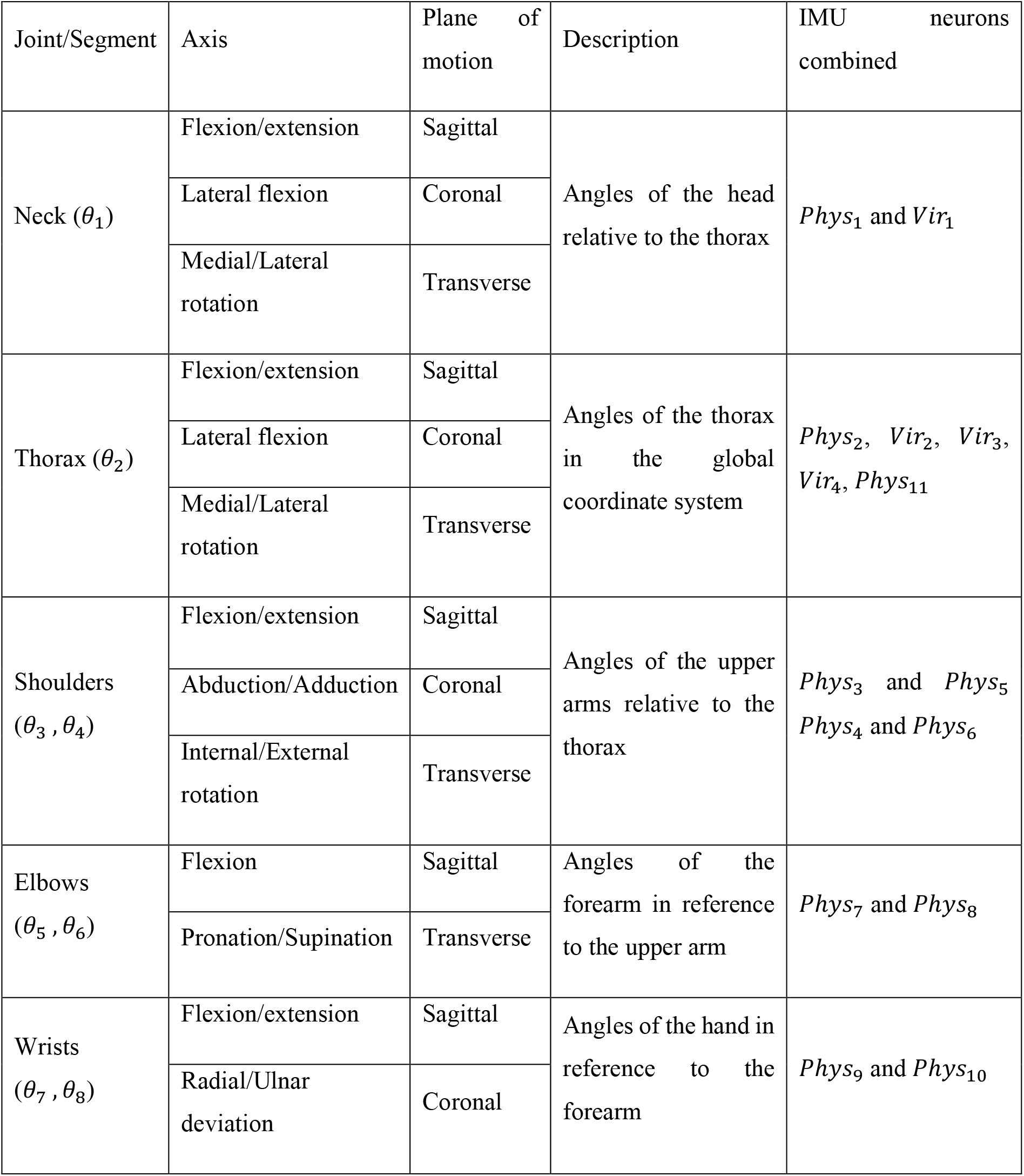
Upper body segments, the subsequent motion considered, reference segment and the neurons involved in the calculation of resultant angles

| Joint/Segment | Axis | Plane of motion | Description | IMU neurons combined |
| --- | --- | --- | --- | --- |
| Neck ($\theta_1$) | Flexion/extension | Sagittal | Angles of the head relative to the thorax | $Phys_1$ and $Vir_1$ |
|  | Lateral flexion | Coronal |  |  |
|  | Medial/Lateral rotation | Transverse |  |  |
| Thorax ($\theta_2$) | Flexion/extension | Sagittal | Angles of the thorax in the global coordinate system | $Phys_2$, $Vir_2$, $Vir_3$, $Vir_4$, $Phys_{11}$ |
|  | Lateral flexion | Coronal |  |  |
|  | Medial/Lateral rotation | Transverse |  |  |
| Shoulders ($\theta_3, \theta_4$) | Flexion/extension | Sagittal | Angles of the upper arms relative to the thorax | $Phys_3$ and $Phys_5$<br>$Phys_4$ and $Phys_6$ |
|  | Abduction/Adduction | Coronal |  |  |
|  | Internal/External rotation | Transverse |  |  |
| Elbows ($\theta_5, \theta_6$) | Flexion | Sagittal | Angles of the forearm in reference to the upper arm | $Phys_7$ and $Phys_8$ |
|  | Pronation/Supination | Transverse |  |  |
| Wrists ($\theta_7, \theta_8$) | Flexion/extension | Sagittal | Angles of the hand in reference to the forearm | $Phys_9$ and $Phys_{10}$ |
|  | Radial/Ulnar deviation | Coronal |  |  |

A laparoscopic trainer (Laparo Aspire, LAPARO Medical Simulators, Wroclaw, Poland) was used to simulate a patient’s abdominal cavity [27]. On the laparoscopic trainer adipose tissue was simulated using foam fixed across the ports. Models of BMI 20, 30, 40 and 50 kg/m^2^ were developed using foam thickness of 1.7 [28], 6.5, 9.5, and 11 cm respectively. Small holes were drilled in the foam allowing the trocars to be placed into the trainer. This study utilised a contralateral port placement and remained the same for all BMI models [19]. The desk height was set at 65 cm for all participants, a height found to be optimal within previous ergonomic studies [29]. During models of BMI 40 and 50 kg/m^2^ a side bar of 7.5 cm in width was connected to the standard operating table to create a wider surface area and replicate the real surgical environment when operating on patients with severe obesity [30,31]. The same laparoscopic training environment and instruments were used by all participants.

### Experimental Protocol

A threading task was selected for the assessment (Figure 2). The task was completed twice for each BMI model of 20, 30, 40 and 50 kg/m^2^. The BMI models were presented in a randomised order with a minimum of three minutes rest taken between trials. The duration of each trial was defined from the moment the participant held the thread with the laparoscopic graspers within the laparoscopic trainer, until the last outer frame of the task had been threaded. No time limit was set during the experiments because fatigue is known to have an effect on posture [1]. Prior to the BMI trials, three familiarisation trials were conducted with no foam on the trainer to minimise any learning bias [33].

**Figure 2.**
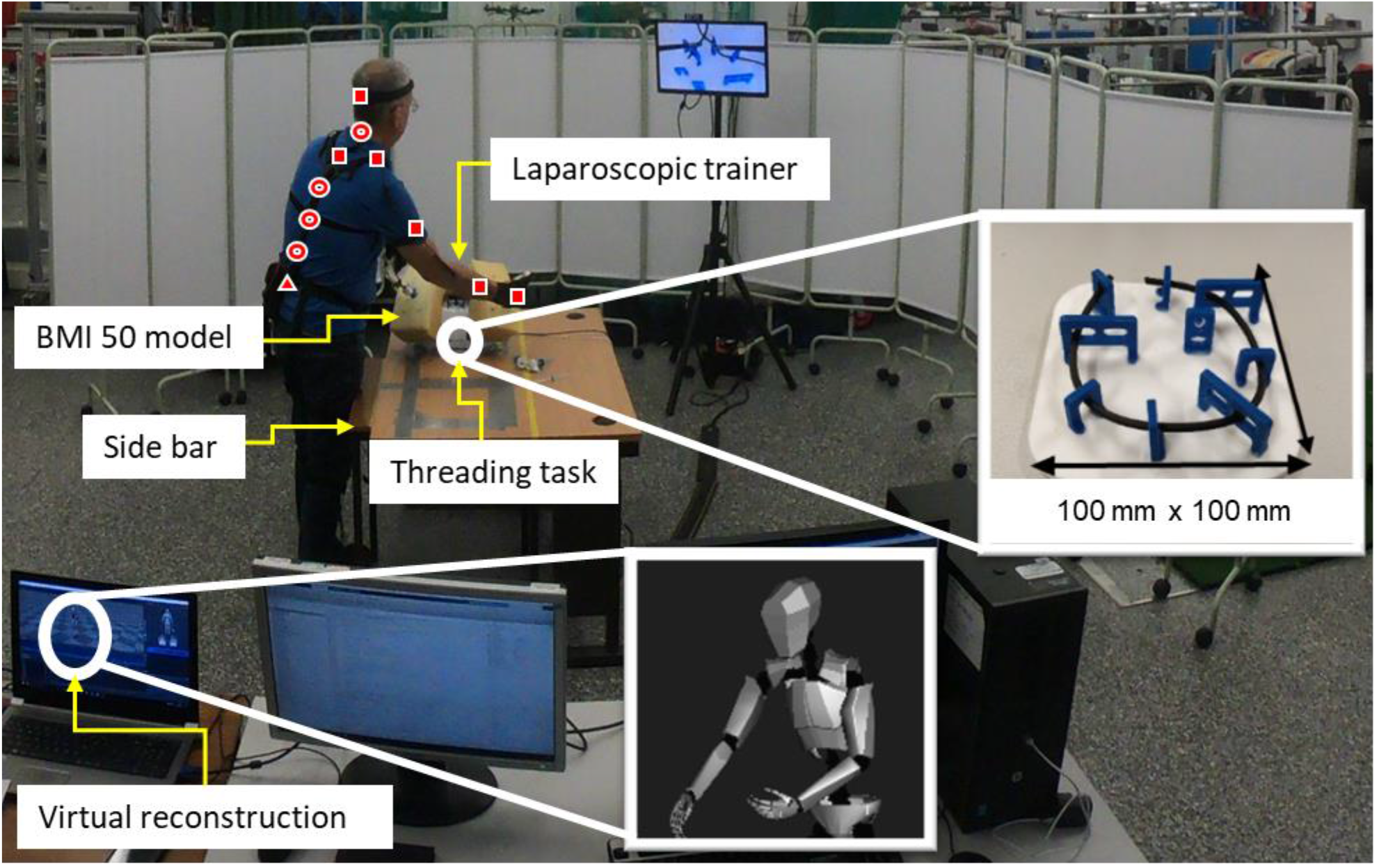
Experimental setup with BMI 50 kg/m^2^ model and Perception Neuron motion capture system described in Figure 1.

### Data preparation

The orientation of the rigid body segments defined in Figure 1B were calculated by following an identical methodology to that described in [23]. To quantify surgeon posture, the human body was modelled as a kinematic system, whereby IMUs were placed on body segments (Figure 1A) to form an interconnected system of links. Rigid body segments or system links (dotted circles in Figure 1B) are connected to one another via joints which facilitate the local expression of segments through child-parent relationships. A joint (shoulder, elbow, wrist) which provides a single point in 3-dimensional space for a segment to rotate about. Thus, a 3-dimensional rotational vector is produced at the joint which represents the amount of angular displacement a child segment experiences in relation to the parent segment in each axis of rotation (Table 2). Therefore, the orientation of each rigid body segment is expressed locally to its parent segment via the conjoining joint. For example, the wrist joint angle represents the amount the hand segment rotates in reference to the forearm. A visualisation of the original kinematic chain has been displayed in Figure 1A, where the root node (lower thorax) is the start of the kinematic chain and is measured in reference to the global coordinate system (*T*_0_) and thereafter each segment is measured in reference to its parent. This behaviour continues until a termination segment (head) or end effector is reached (hands).

To obtain joint angles expressed relative to the appropriate parent segment and in a format suitable for ergonomic assessment, (i.e. head in reference to the thorax) the default joint angle outputs from the Perception Neuron model required modifying. In addition, the Perception Neuron motion capture system produces orientation outputs for each neuron in the format of quaternions. Quaternions express a 3D rotation as a 4D complex vector and provide a more efficient and reliable methodology of representing orientation that is not susceptible to edge case issues such as gimbal lock [34]. The motion capture system provides quaternion outputs for each neuron in Figure 1A in the local coordinate system. In order to utilise the suits default outputs in a manner that would reference body segments to one another by the appropriate anatomical joint (Figure 1B), quaternion outputs were combined (Table 2). Due to the superior computational efficiency of quaternions, quaternion multiplication was used to combine the quaternions and therefore rotations of specific sets of Figure 1A neurons. Once combined, the resultant quaternions were converted to Euler angles to provide joint rotations for child segments in reference to their parent in a format suitable for ergonomic assessment.

By default, the Perception Neuron model produces a quaternion outputs, *N*_2_, for the head neuron (*phys*_1_) relative to the virtual neck neuron (*Vir*_1_) and, *N*_1_, for the virtual neck neuron relative to the C7 neuron (*phys*_2_) (Figure 1A). The orientation of *phys*_1_ and *Vir*_1_ neuron rotations were combined using quaternion multiplication in order to treat both neurons as one rigid body segment in reference to the thorax (Figure 1B). Once combined, the resultant quaternion vector (*θ*_1_) represented the orientation of the head rigid body segment when defined at the neck joint (Figure 1B) in reference to the thorax. The orientation of the thorax rigid body segment in the global coordinate system (*θ*_2_, Figure 1B) was obtained by combining the quaternion outputs (*T*_0−4_, Figure 1A) from the respective thorax neurons (*phys*_11_, *Vir*_4_, *Vir*_3_, *Vir*_2_, *phys*_2_, Figure 1A). The orientation of the upper arm rigid body segments were obtained by combining the quaternion outputs (*LS*_1−2_ and *RS*_1−2_, Figure 1A) from their respective upper arm neurons (*phys*_3_, *phys*_5_ and *phys*_4_, *phys*_6_) when defined at their respective shoulder joints (*θ*_3_, and *θ*_4_, Figure 1B) in reference to the thorax. The orientation of the hands and forearms did not require any modification because they are by default defined at the wrists and elbows and therefore, expressed in reference to the forearm and shoulder segments, respectively. This computational methodology alongside the combination of the specified neuron sets to obtain the joint/segment angles for the upper body, have been previously compared to a gold standard in optoelectronic motion capture and demonstrated sufficient measurement precision (3 – 5°) for this application [23].

### Postural assessment

The LUBA ergonomic framework was used to assess the motion capture data. This framework was selected because it is upper body focused and has been shown to be most effective in medium-risk conditions [35]. An emphasis was placed on the use of a framework suited to medium-risk circumstances because LS has been shown to induce prolonged low-risk non-neutral postures as well as episodic high-risk postures [10,36,37], therefore a medium risk classification seemed to be the optimal choice given these postural habits.

The LUBA framework was developed based on perceived discomfort scores provided by participants who completed functional movements in each axis of rotation. Discomfort scores were associated with dichotomous movement range categories for each axis of rotation through perceived feedback from study participants [38]. In general, the closer to the maximum range of motion for a given rotation, the larger the LUBA score. Hence, non-neutral postures are scored higher and larger LUBA scores represent higher risk postures. A MATLAB script was written to implement the LUBA dichotomous movement range categories for each segment and joint axis of rotation. Specifically, the relative discomfort scores as defined by the LUBA framework in standing postures were used to classify the posture of each joint and segment [38]. The pre-processed Euler angle data for each joint (neck, shoulders, elbows, wrists) and segment (thorax) was then streamed into the script and classified into the appropriate categories producing sub-scores for each component. The LUBA sub-scores were then combined using Equation 1 to produce the final overall LUBA posture score. The overall LUBA scores were then grouped based on surgeon experience level and the BMI level tested.

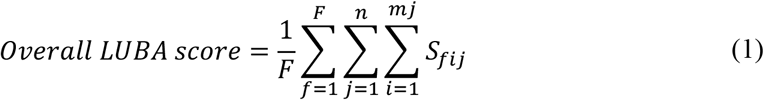

where *i* is the joint/segment angle counter, *j* is the joint/segment counter; *n* is the total number of joints/segments included, m_*j*_ is the total number of joint/segment angles included for the *j*th joint/segment, *f* is the time frame counter, *F* is the total number of time frames, and *S*_*fij*_ is the relative discomfort score of the *i*th joint/segment angle in the *j*th joint/segment at an *f*th instance in time [38]. Once an overall LUBA score was calculated for the entire upper body it was sorted into one of four posture categories defined in [38]. However, the previously defined overall LUBA posture categories were modified to accommodate the assessment of both upper extremities, as the predefined categories were developed when only considering one upper extremity. Therefore, category 1 covers postures with an overall LUBA score ≤ 10, category 2 covers postures from 10 < overall LUBA score ≤ 20, category 3 covers postures from 20 < overall LUBA score ≤ 30 and category 4 covers postures with an overall LUBA score > 30. The LUBA corrective actions which correspond to each posture category have been included. While these actions do not correspond to firm timings and detailed conditions, they do provide context for the overall LUBA score. Category 1 advises no corrective actions, category 2 suggests further investigation and corrective changes without immediate intervention, category 3 requires gradual corrective action and category 4 requires immediate intervention and corrective action.

In conjunction with overall LUBA scores described previously, an additional normalised overall LUBA score was calculated to isolate the effect of patient BMI on surgeon posture. The normalised overall LUBA score was taken by normalising overall LUBA scores for the obese and severely obese BMI models for each participant (BMI 30 – 50 kg/m^2^) to the overall LUBA scores produced by their performance on the baseline BMI 20 kg/m^2^ model. These normalised results were then sorted into the relevant experience groups and averaged to give a result for each group. In theory, this method will reduce the variability of the overall LUBA scores within each group and emphasize the effect of BMI on overall LUBA score.

### Task completion time

The total time (in seconds) to complete the laparoscopic task was also evaluated to provide context for the overall LUBA scores (Table 3). The task completion time was defined as the duration of time to successfully complete the threading task as specified within the experimental protocol.

**Table 3.** Mean (± Standard error) task completion time (s)

| Experience group | BMI (kg/m<sup>2</sup>) |  |  |  |
| --- | --- | --- | --- | --- |
|  | 20 | 30 | 40 | 50 |
| Novices | 457 ($\pm 216$) | 465 ($\pm 238$) | 601 ($\pm 359$) | 614 ($\pm 288$) |
| Intermediates | 233 ($\pm 60$) | 201 ($\pm 26$) | 284 ($\pm 101$) | 337 ($\pm 74$) |
| Experts | 214 ($\pm 24$) | 215 ($\pm 30$) | 214 ($\pm 22$) | 276 ($\pm 42$) |

### Statistical analysis

All statistical analysis was conducted in SPSS (IBM SPSS Statistics, v. 24, IBM Corp., Armonk, NY). Mean responses for overall LUBA score, normalised overall LUBA score and task completion times were taken by grouping the participant data by experience level and averaging for each BMI level. Two-way mixed analysis of variance (ANOVA) was conducted with input categories of experience and BMI level for LUBA score and task completion time. Also, a one-way ANOVA was conducted with an input of normalised LUBA score, as the experience group factor is inherently excluded due to the normalisation process. Where significant main effects were found, post hoc tests were carried out using least significant difference tests. An alpha level of α ≤ 0.05 was set for all statistical testing. Effect sizes were calculated as the partial eta squared value (η_*p*_); small effect size (η_*p*_ = .0099), medium effect (η_*p*_ = .0588) and large effect size (η_*p*_ = .1379) [39].

## Results

Participants completed the task twice at each BMI level, except for one novice, who finished the task once at each BMI level due to long task completion times which prevented any repeats in the allocated testing session.

### Postural assessment

There were no significant interaction effects between experience group and BMI level on LUBA score (p = 0.316, η_*p*_ = 0.265). There were significant main effects of BMI on overall LUBA scores (p = 0.021, η_*p*_ = 0.364) and normalised overall LUBA scores (p = 0.035, η_*p*_ = 0.209). There was also a significant main effect of experience group on overall LUBA score (p = 0.05, η_*p*_ = 0.576). Post hoc tests were performed on both the overall LUBA score and normalised overall LUBA score. The following post hoc results correspond to the effect of BMI level on the normalised overall LUBA score due to the isolation of obesity effects and the effect of experience level of overall LUBA score.

The results from the post hoc tests on the effect of BMI level on normalised overall LUBA scores showed significant increases during BMI 50 kg/m^2^ as compared to BMI 20 kg/m^2^ and 30 kg/m^2^ (p = 0.15 and p = 0.20 respectively), with the remainder of the results showing non-significant differences. The results from the post hoc tests on the effect of experience group on overall LUBA scores revealed significant differences for novices compared to experts (p = 0.02), with the other comparisons yielding non-significant results.

Novice participants produced the most severe LUBA scores at each BMI level, with overall LUBA scores for all obese models > 25 (Category 3, gradual corrective actions required). The intermediate group also exhibited large overall LUBA scores of > 20 when subject to obese models (Category 3). Expert overall LUBA scores were the lowest across all BMI levels displaying scores of 10 < overall LUBA score ≤ 20 (Category 2, however corrective actions are still suggested). In general, the less experienced surgeons exhibited larger overall LUBA scores at all BMI levels as compared to the more experienced surgeons and larger BMI levels increased overall LUBA scores within each experience group.

Within all experience groups and BMI levels, the major contributor to overall LUBA scores was the thorax. The higher BMI models increased the thorax LUBA score more so than any other (Figure 4). Moreover, the most substantial contributor to the thorax LUBA scores was sub-optimal thorax lateral flexion with scores ranging from 5.0 – 6.3 for novices, 3.2 – 5.3 for intermediates and 1.9 – 3.6 for experts across BMIs 20 kg/m^2^ to 50 kg/m^2^ (Figure 5). In all experience groups, the LUBA scores for both wrists increased when exposed to severely obese BMI models, indicating a degradation in posture for both hands. Moreover, novice and intermediate right upper arm and right forearm posture worsened with increasing BMI level, whereas experts left forearm posture appeared to worsen with increasing BMI.

**Figure 4.**
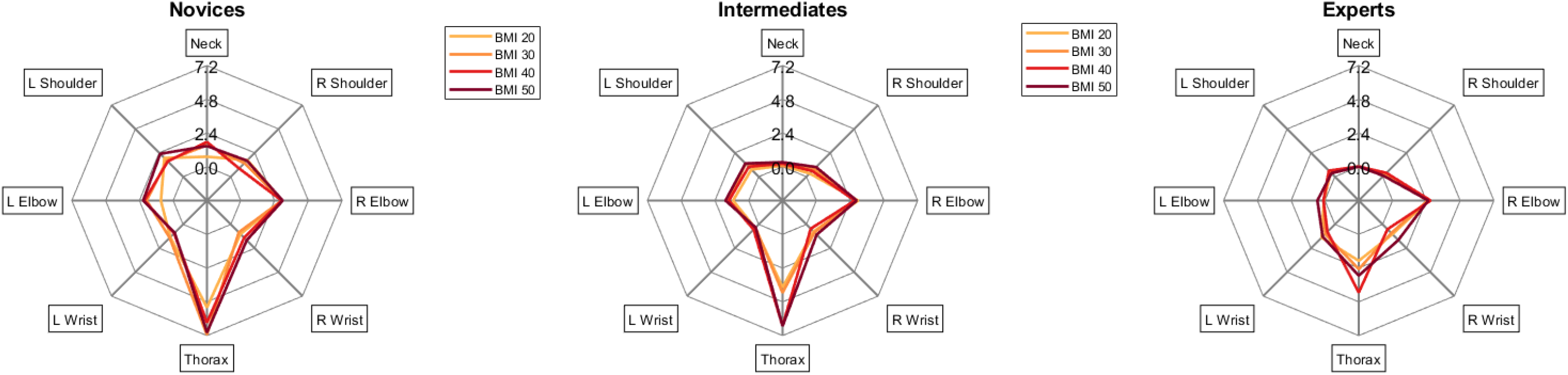
The breakdown of the overall LUBA score into mean individual joint/segment LUBA scores for novice, intermdiate and experts at each BMI level

**Figure 5.**
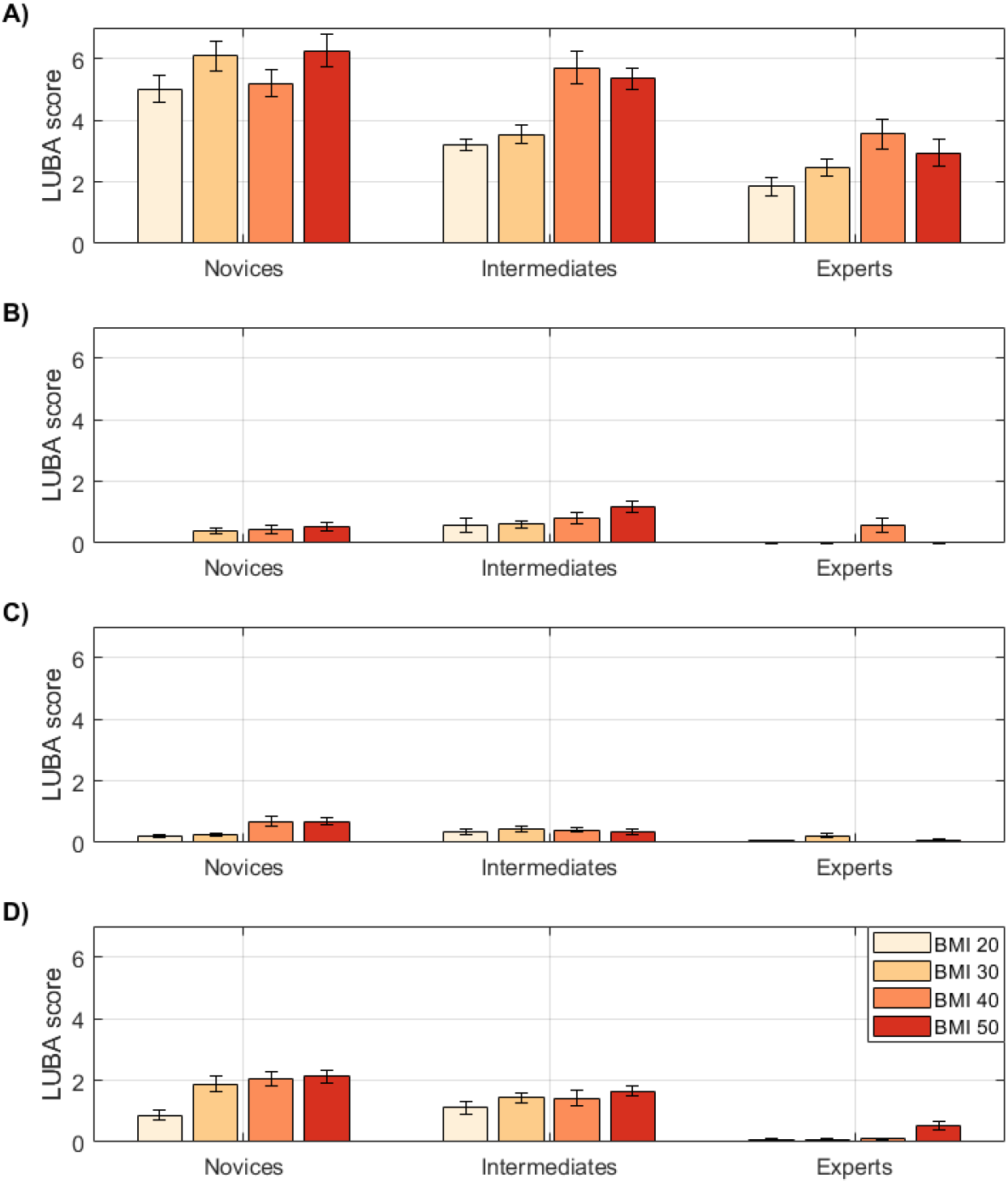
Mean (± Standard error) LUBA scores for (A) Thorax lateral flexion, (B) Thorax axial rotation, (C) Left shoulder abduction/adduction and (D) Right shoulder abduction/adduction for the novice, intermediate and expert surgeons at each BMI level

### Task completion times

There was no significant interaction effect between experience group and BMI level (ρ = 0.663, η_*p*_ = 0.124) on task completion times. There was no significant main effect of experience level (ρ = 0.507, η_*p*_ = 0.177) or BMI (ρ = 0.101, η_*p*_ = 0.318) on task completion time (Table 3), however large effects sizes were observed. Novices took the most time to complete the simulated task at each BMI level with substantial variability in completion time within the group. In general, the time to complete the task progressively increased with BMI, which means that the largest LUBA scores were maintained for the longest durations.

## Discussion

Musculoskeletal disorders amongst laparoscopic surgeons and the wider surgical community are well recognised, with the adoption of non-neutral postures during surgery often attributed as a major task-related cause [7,10,11,37,40]. Surgeons often perceive patients with obesity to be an aggravator of this task-related cause [13,17]; however, there remains little objective analysis to support this. The purpose of this study was to objectively quantify the effect of different levels of simulated patients with obesity (30 kg/m^2^) and severe obesity (40 – 50 kg/m^2^) on the posture of laparoscopic surgeons with varying levels of surgical experience. The upper body posture of surgeons was assessed by classifying motion capture data using the LUBA ergonomic framework. This study contributes to the growing literature concerning the impact of patient BMI on surgeons performing laparoscopy [9,13,17,19,20].

BMI 50 kg/m^2^ caused significant deterioration in postural neutrality (i.e. increased overall LUBA score) as compared to 20 kg/m^2^ and 30 kg/m^2^ in the participant cohort (Figure 3). Moreover, expert surgeons suffered substantial degradation in overall LUBA scores during severely obese models as compared to the baseline (20 kg/m^2^), which emphasises the negative effect that severe obesity models had on the postural performance of surgeons regardless of previous experience or expertise. All groups suffered postural deterioration with increasing BMI level, however novice differences varied considerably in comparison to the other groups. The ability of novice surgeons to minimise non-neutral postures was highly variable, a similar result has been reported previously [20]. The novice group sustained all sub-optimal postures for much larger durations as compared to the other experience groups due to larger LUBA scores and task completion times (Figure 4, Table 3). Specifically, at BMI 50 kg/m^2^ where the highest LUBA scores and longest trial times were recorded, on average, novice overall LUBA scores were approximately 25% larger and double those recorded for the intermediates and experts, respectively. In addition, novice trial durations lasted approximately double the amount of time of both the more experienced groups. All groups recorded larger overall LUBA scores for longer durations as the BMI models increased, which ultimately increased the postural workload and the burden on the musculoskeletal system, more so for the novices than the other experience groups.

**Figure 3.**
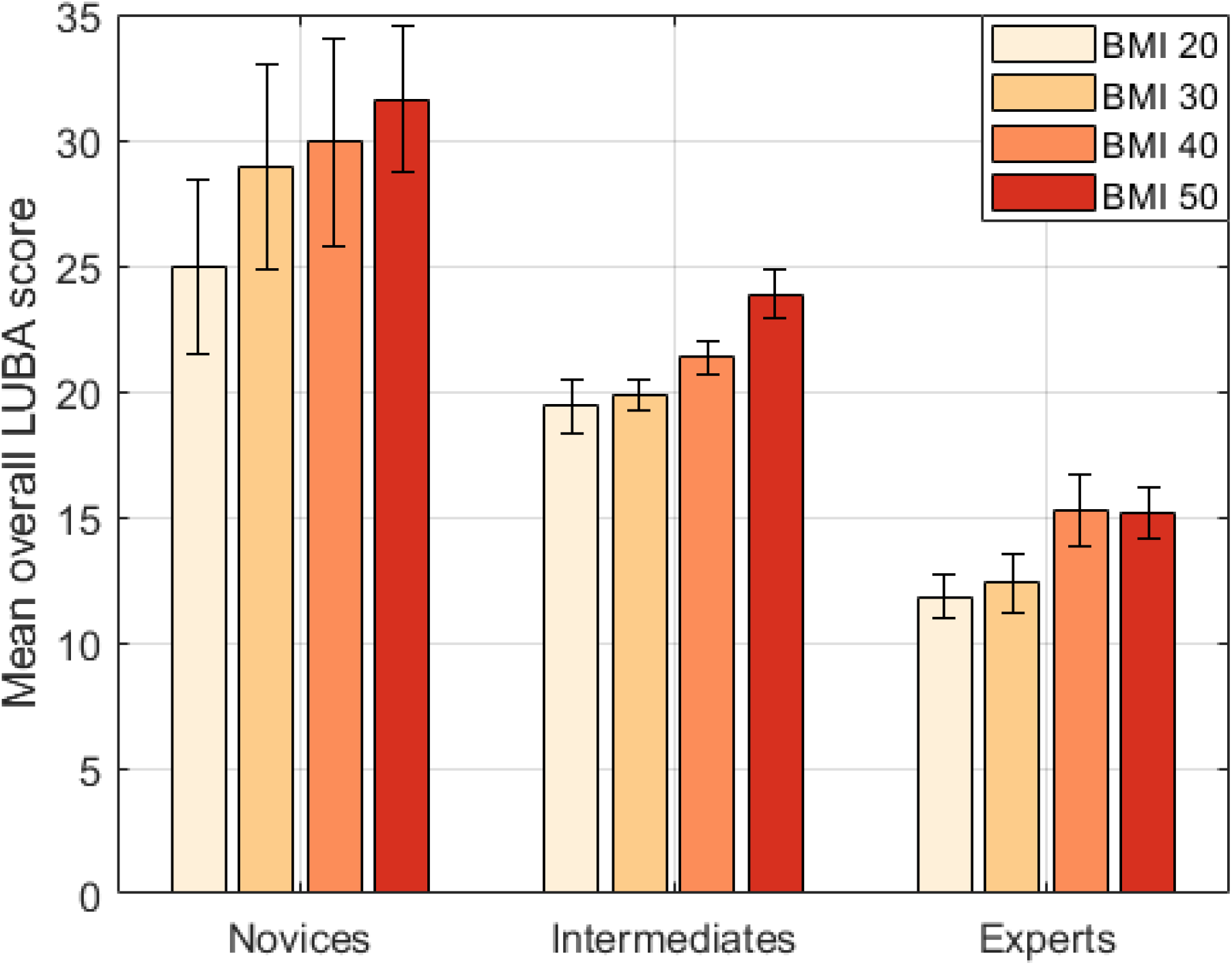
Mean (± Standard error) novice, intermediate and expert LUBA score

The anatomical area that displayed the largest LUBA score across all groups was the thorax (Figure 4). In theory, the thorax is an area where optimisation to minimise LUBA score is feasible as the segment is constrained by passive variables such as port placement, table position and patient BMI. Large thorax scores appeared to be systemic across all BMI and experience levels, however scores increased markedly when attempting severely obese BMI models and this was principally linked to degradation in lateral flexion posture (Figure 5A). The increase in BMI level and addition of the side bar induced more severe thorax lateral flexion for all groups, particularly intermediates and experts whose lateral flexion LUBA scores increased by 70% and 90% respectively when compared to the baseline model. The novice LUBA score for the thorax was the largest of all groups at each BMI level, however no substantial increase occurred, and the score remained large regardless of BMI.

Excessive thorax axial rotation and shoulder abduction (Figure 5B, 5C, 5D) in novice and intermediate surgeons are also postures which could be improved given that experts showed very little incidence of poor posture in these areas regardless of BMI level. The appropriate manipulation of the joint contributions (shoulder, elbow, wrist) within the upper extremity can allow for an infinite number of redundant motion patterns to achieve the same outcome [41,42]. As the upper arm segment is further up the kinematic chain and does not directly influence instrument mechanics, reducing sub-optimal shoulder conditions is more feasible allowing the elbow and wrist joints to take the burden of the procedure. This kinematic profile was exhibited by the expert group as shoulder LUBA scores were close to zero with larger LUBA scores for the elbows and wrists as compared to the less experienced groups. Thus, this kinematic approach could form the basis for future training and optimisation targets of less experienced surgeons in order to reduce the sub-optimal impact on the shoulder joints.

The LUBA framework identified the thorax, the right elbow, and both wrists as the segments/joints with the largest contributors to the overall LUBA score (Figure 4). Approximate, linear relationships were found for both experience level and BMI level in reference to LUBA scores; for these segments/joints increasing the model BMI worsened posture and more experienced surgeons had better posture. The identification of wrists and elbows as large contributors to overall LUBA score is insightful, however optimisation in these areas may be more challenging than areas further up the kinematic chain [41]. Indeed, every procedure will have individual nuances, therefore no scenario can effectively ensure neutral hand and forearm postures for set tasks as task requirements are intrinsically chaotic. Therefore, as laparoscopic hand and forearm posture is largely dictated by the required instrument manipulation inherent of a given task, the pursuit for safer postures may compromise task performance. Thus, it is reasonable to suggest that any targeted optimisation of wrist and elbow posture would need to be considered carefully to ensure task performance was not compromised, especially when similar behaviour was exhibited by the experts.

The lack of extensive experience in a laparoscopic environment and exposure to patients with obesity are the likely causes of poor posture for the novice surgeons. In addition, the contralateral port placement is an ecological factor that may have promoted excessive thorax lateral flexion to access the port site on the right side, especially when using severely obese models. Port site selection is a highly subjective decision amongst laparoscopists, with many factors such as patient BMI, pathology and dominant hand influencing a surgeon’s decision [19]. The contralateral port placement setup was chosen over a midline or ipsilateral setup due to its preference amongst early career surgeons (less than 5 years’ experience) and its recognized risk to the postural wellbeing of surgeons [19]. Thus, selecting a contralateral port setup enabled the simulation of high-risk scenarios that young surgeons (less than 5 years’ experience) may find themselves in early on in their careers.

The key findings of this study were that performing LS on the higher BMI models significantly deteriorated the upper body posture of novice, intermediate and expert surgeons. In addition, the less experience a surgeon has, the worse their posture is likely to be. Larger LUBA scores translate to an increased risk of MSDs [35,38], which implies that larger BMI models and less surgical experience increases the risk of MSDs in surgeons. Also, the thorax and dominant shoulder have been identified as anatomical areas with the most severe LUBA scores that have feasible scope for improvement. Novice participants are not usually exposed to patients with obesity while developing their laparoscopic ability, however, early career exposure to obese models could enable biomechanical optimisation of the surgeon while also ensuring laparoscopic task proficiency. This could enable a reduction in unnecessary non-neutral postures and poor postural habits in simulated obese environments, thus reducing the incidence of these factors and risk of MSDs in live scenarios.

There are limitations that need to be recognised when interpreting the results from this study. The small sample size, completion of a single task and implementation of one port placement may reduce the significance of the relationship between the surgeon experience, BMI level and LUBA scores. In particular, the ANOVA results for task completion times (low non-significant p values and large effects sizes) as well as the results pertaining to the non-significant BMI effects suggest that the small sample sizes resulted in lower statistical power and inhibited the ability to detect significant differences in those cases. Thus, it would be expected that larger sample sizes would increase the statistical power leading to significant differences in both the aforementioned results. From the statistical data related to the task completion times, the sample size required to produce significant main effects was estimated as 30 (10 in each experience group) [43].

An additional potential limitation is that the use of the LUBA ergonomic framework may overlook lower-risk postures and subsequently underestimate the incidence of non-neutral posture. Therefore, the outcome of the postural assessment is undoubtedly sensitive to the framework applied. Finally, the ecological validity of using foam to simulate body fat may be seen to be unrepresentative, however, feedback to support its use has been reported [20].

This study quantified the posture of surgeons by applying a segment/joint angle focussed analysis technique. A future assessment that considers segment/joint angles in conjunction with other biomechanical aspects of performance (e.g. both kinematics and kinetics), cumulative time in non-neutral postures and the total work involved in procedures should be investigated at different BMI levels.

## Conclusion

Conducting LS on simulated severely obese increases the duration and severity of non-neutral postures as compared to LS on normal BMI models. In addition, surgeons with less surgical experience adopt more extreme non-neutral postures across all BMI models, suggesting that early career biomechanical optimisation could benefit this group to minimise the incidence of extreme poor postural habits. The increase in average time spent in severe non-neutral postures when exposed to obese models is more physically demanding and aggravates the musculoskeletal workload of the surgeon, which ultimately increases their risk of MSDs. Strict management of workloads, including exposure to patients with severe obesity may be necessary to reduce the risk of MSDs in surgeons.

## Data Availability

Restriction on data availability until publication and the inherent embargo period has elapsed

